# Open-Source Offline-Deployable Retrieval-Augmented Large Language Model for Assisting Pancreatic Cancer Staging

**DOI:** 10.64898/2025.12.26.25343050

**Authors:** Hisashi Johno, Akitomo Amakawa, Atsushi Komaba, Ryota Tozuka, Yuki Johno, Junichi Sato, Kentaro Yoshimura, Kazunori Nakamoto, Shintaro Ichikawa

## Abstract

**Purpose:** Large language models (LLMs) are increasingly applied in radiology, but key challenges remain, including data leakage from cloud-based systems, false outputs, and limited reasoning transparency. This study aimed to develop an open-source, offline-deployable retrieval-augmented LLM (RA-LLM) system in which local execution prevents data leakage and retrieval-augmented generation (RAG) improves output accuracy and transparency using reliable external knowledge (REK), demonstrated in pancreatic cancer staging.

**Materials and Methods:** Llama-3.2 11B and Gemma-3 27B were used as local LLMs, and GPT-4o mini served as a cloud-based comparator. The Japanese pancreatic cancer guideline served as REK. Relevant REK excerpts were retrieved to generate retrieval-augmented responses. System performance, including classification accuracy, retrieval metrics, and execution time, was evaluated on 100 simulated pancreatic cancer CT cases, with non-RAG LLMs as baselines. McNemar tests were applied to TNM staging and resectability classification.

**Results:** RAG improved TNM staging accuracy for all LLMs (GPT-4o mini 61%→90%, *p*<0.001; Llama-3.2 11B 53%→72%, *p*<0.001; Gemma-3 27B 59%→87%, *p*<0.001) and mildly improved resectability classification (72%→84%, *p*=0.012; 58%→73%, *p*=0.006; 77%→86%, *p*=0.093), with Gemma-3 27B showing performance comparable to GPT-4o mini. Retrieval performance was high (context recall = 1; context precision = 0.5–1), and local models ran at speeds comparable to the cloud-based GPT-4o mini.

**Conclusion:** We developed an offline-deployable RA-LLM system for pancreatic cancer staging and publicly released its full source code. RA-LLMs outperformed baseline LLMs, and the offline-capable Gemma-3 27B performed comparably to the widely used cloud-based GPT-4o mini.

## Introduction

Large language models (LLMs) hold promise for transforming radiology by assisting across a variety of tasks, including patient record summarization, medical decision-making, report generation, research data management, and trainee education (1). However, their safe and effective clinical implementation will require overcoming several key limitations, such as the generation of false information (so-called hallucinations), limited transparency and reliability in reasoning, and inherent risks to data privacy and security (1). Retrieval-augmented generation (RAG), which integrates domain-specific knowledge into LLM queries, has recently emerged as a promising approach in radiology to mitigate hallucinations and enhance the transparency and traceability of LLM outputs (2). In addition, compared with proprietary cloud-based LLMs, open-source LLMs generally exhibit lower performance but offer advantages in data privacy and security by enabling local execution (3). Taken together, further development of high-performance, offline-deployable retrieval-augmented large language models (RA-LLMs) is warranted in radiology.

In radiology, most evidence supporting the usefulness of RAG has come from studies using cloud-based LLMs (2–6), but reports on offline-deployable RA-LLMs remain extremely limited. Wada et al. recently demonstrated that RAG improves the performance of a local LLM (Llama 3.2 11B) for radiology contrast media consultation. However, the embedding model used for RAG (text-embedding-3-large) is cloud-based, meaning that the overall system cannot be operated entirely offline. Furthermore, the source code for system construction has not been made publicly available (7). Choi et al. developed a locally executable RA-LLM (base LLM: Llama-3 7B; embedding model: paraphrase-multilingual-MiniLM-L12-v2) for PET imaging reports, demonstrating its utility in differential diagnosis, although the source code was not released (8). Similarly, Welsh et al. built a local RA-LLM (base LLM: mistral-7b-instruct-v0.2; embedding model: intfloat/e5-mistral-7b-instruct) for radiology research assistance, but its source code was likewise unavailable (9). Note that, in radiology, sharing and accumulating open-source code are increasingly regarded as vital for ensuring transparency and reproducibility and for advancing artificial intelligence research (10). As a study that released its source code, Weinert et al. demonstrated that RAG enhances the response accuracy of LLMs in radiology examinations. Among the LLMs used, Command R+ and Mixtral are locally executable; however, the embedding model used for RAG (text-embedding-3-large) is cloud-based, therefore the overall system is not offline-deployable (11). Taken together, truly open-source, offline-deployable RA-LLM implementations in radiology remain scarce.

This study aimed to develop an offline-deployable RA-LLM system for diagnostic radiology and to release it as open-source code. Cancer staging is one of the essential and complex tasks in diagnostic radiology, with pancreatic cancer staging being particularly challenging. Therefore, we evaluated the performance of the proposed RA-LLM system in pancreatic cancer staging using 100 simulated CT cases.

## Materials and Methods

### System Architecture

The proposed RA-LLM system was implemented in Python 3.13. Three LLMs were used as the base models: Llama-3.2 11B (Meta), Gemma-3 27B (Google), and GPT-4o mini (OpenAI). When local models (Llama-3.2 11B or Gemma-3 27B) were used, the entire system was capable of fully offline operation, whereas GPT-4o mini served as a representative cloud-based model for comparison. The overall system configuration is illustrated in Figure 1 and is described in detail below.

**Figure 1:**
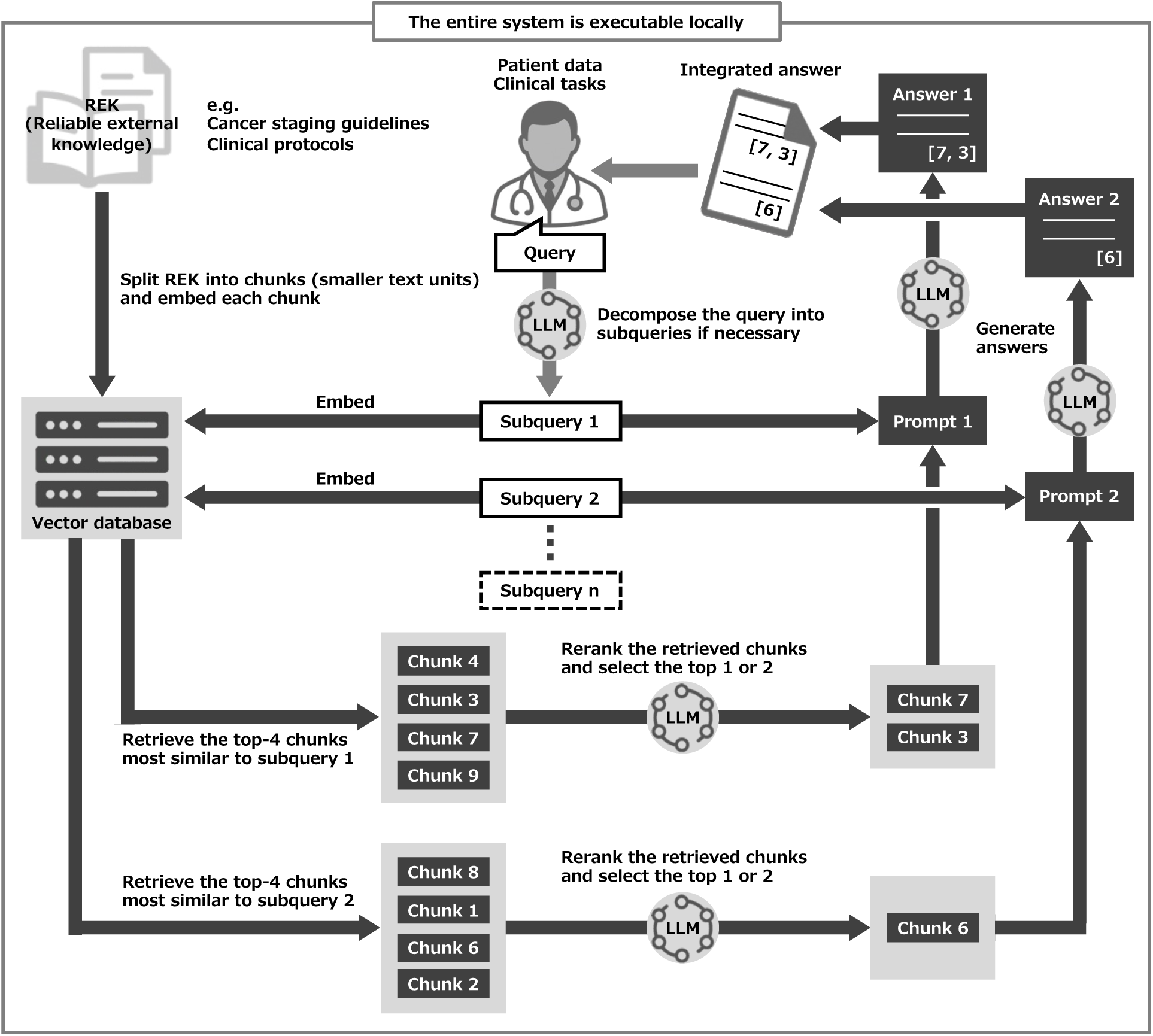
System architecture of our offline-deployable retrieval-augmented LLM (RA-LLM). A clinician-written query, based on patient data and clinical tasks, is decomposed into subqueries by a specified LLM. For each subquery, the top four similar chunks (smaller text units) are retrieved from the reliable external knowledge (REK) and then reranked by the LLM to select the top one or two. These selected chunks are combined with the subquery to form the prompt, and the LLM generates an output while retaining the IDs of the referenced chunks. Thereafter, the subquery-level outputs are integrated into a single answer, which is further reformatted into a structured final output. Note: All LLM-invoked processes in this workflow use the same LLM. LLM = large language model.

As the reliable external knowledge (REK) to be referenced, we used the full text (in Markdown format) of the *Eighth Edition of the Japanese Classification of Pancreatic Carcinoma* (12). The REK was divided into multiple chunks (smaller text units) using a standard Markdown-based text splitting method, with a chunk size of 1500 tokens and a chunk overlap of 500 tokens. Each chunk was embedded with the BAAI/bge-base-en-v1.5 model and stored locally in a FAISS vector database.

Given a user query consisting of patient data and clinical tasks (Figure 2), a specified LLM (GPT-4o mini, Llama-3.2 11B, or Gemma-3 27B) automatically decomposed the query into multiple subqueries when appropriate. Each subquery was vectorized by the embedding model, and the four most similar chunks were retrieved based on Euclidean distance. These retrieved chunks were then reranked by the LLM according to their relevance to the subquery, and the top one or two chunks were selected. The selected chunks were combined with the subquery to create the prompt, and the LLM generated an output while preserving the IDs of the referenced chunks. Thereafter, the subquery-level outputs were integrated into a single answer, which was further reformatted into a structured final output.

**Figure 2:**
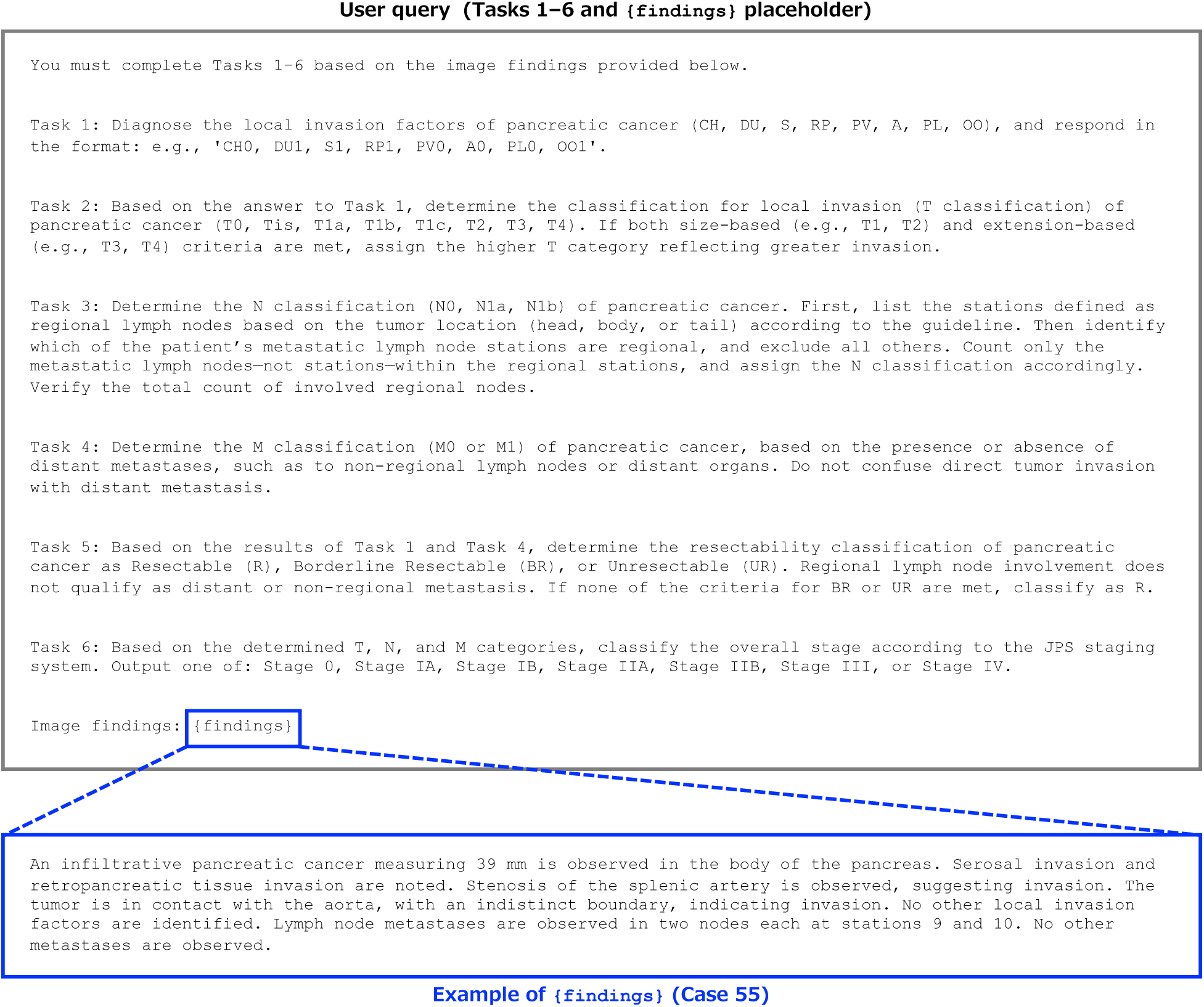
User query template with example image findings (Case 55). For each pancreatic cancer case, a query was constructed based on CT findings to request the classification of local invasion (Task 1), T category (Task 2), N category (Task 3), M category (Task 4), resectability (Task 5), and overall TNM stage (Task 6) for pancreatic cancer.

All experiments were conducted on a PC running Windows 11 Home. The workstation was equipped with an AMD Ryzen 9 7900 processor, 64 GB of RAM, a 1 TB SSD, and an NVIDIA GeForce RTX 4090 GPU with 24 GB of VRAM. The NVIDIA graphics driver (version 560.94) was used throughout the experiments.

### Pancreatic Cancer Dataset

To evaluate system performance, we used 100 simulated pancreatic cancer cases with CT findings described in English, constructed by Japanese radiologists and previously used to assess the cloud-based RA-LLM system, NotebookLM (5). The dataset appears to reflect the typical stage imbalance encountered in clinical practice, while also including a small number of rare cases so that all staging components are represented.

### Code and Data Availability

The complete system code and all raw experimental data are publicly available in the following repository: https://github.com/mohehe1234/local-rag/tree/v1.0.0-with-results

### Data Analysis

In response to a user query requesting pancreatic cancer staging (Figure 2), our system produced a structured output for each staging component as the final answer: local invasion (CH0 or CH1, DU0 or DU1, S0 or S1, RP0 or RP1, PV0 or PV1, A0 or A1, PL0 or PL1, and OO0 or OO1), T category (T0, Tis, T1a, T1b, T1c, T2, T3, or T4), N category (N0, N1a, or N1b), M category (M0 or M1), resectability (R, BR, or UR), and TNM stage (Stage 0, Stage IA, Stage IB, Stage IIA, Stage IIB, Stage III, or Stage IV). The correctness of these outputs was evaluated over the 100 cases. For local invasion, a case was counted as correct only when all eight factors (CH, DU, S, RP, PV, A, PL, and OO) were correctly classified. For TNM stage and resectability, the exact McNemar test was applied for each base LLM (GPT-4o mini, Llama-3.2 11B, or Gemma-3 27B) to test the null hypothesis that the population proportions of correctly classified cases were equal between the groups with RAG and without RAG, and the corresponding *p* values were calculated.

In addition to the classification accuracy, we evaluated the retrieval performance of our RA-LLM system based on the RAGAS framework, particularly using context recall and context precision (13,14). Let *A* denote the set of all REK chunks and 2*^A^* its power set. For each staging task (local invasion, T category, N category, M category, resectability, or TNM stage), define a function *f*: 2*^A^* → {0,1} by

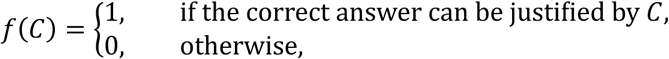

and let *c*_1_, *c*_2_, …, *c*_3_ ∈ *A* be the retrieved chunks in ranked order. The definition of *f* was independently reviewed by two radiologists and one gastroenterologist, and consensus was achieved. As expected, *f* satisfied the following properties: *f*(*A*) = 1, *f*(∅) = 0, and *f*(*C*) = 1 whenever *f*(*C*^’^) = 1 for some *C*^’^ ⊂ *C*. The context recall is given by

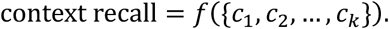

For context precision, each retrieved chunk is labeled as relevant or not. We regard a chunk *c* ∈ *A* as relevant if *f*({*c*}) = 1 and irrelevant if *f*({*c*}) = 0. Then, the context precision is given by

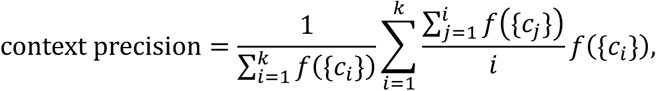

provided that *f*({*c_i_*}) = 1 for some *i*. This definition can be insufficient when an answer requires combining multiple chunks, for example when *f*({*c_i_*}) = 0 for all *i* but *f*N{*c_i_*, c*_j_*}O = 1 for some *i* ≠ *j*. However, such situations did not occur in our experiments. Stated informally, context recall indicates whether the retrieved information is sufficient to justify the correct answer, whereas context precision reflects how effectively the retriever places the required information near the top of the ranked chunks.

For each LLM (GPT-4o mini, Llama-3.2 11B, or Gemma-3 27B), with or without RAG, we measured the duration (in seconds) required for our system to perform pancreatic cancer staging per case. The median and interquartile range (IQR) across the 100 cases were computed, and the results were visualized using box plots.

## Results

As shown in Figure 3, TNM staging accuracy improved markedly with RAG across all LLMs: GPT-4o mini (61%→90%, *p*<0.001), Llama-3.2 11B (53%→72%, *p*<0.001), and Gemma-3 27B (59%→87%, *p*<0.001). For resectability classification, RAG also improved accuracy, albeit more modestly: GPT-4o mini (72%→84%, *p*=0.012), Llama-3.2 11B (58%→73%, *p*=0.006), and Gemma-3 27B (77%→86%, *p*=0.093). Notably, GPT-4o mini and Gemma-3 27B showed comparable performance. Similar trends were also observed for the T, N, and M categories and for local invasion, as shown in Table 1.

**Figure 3:**
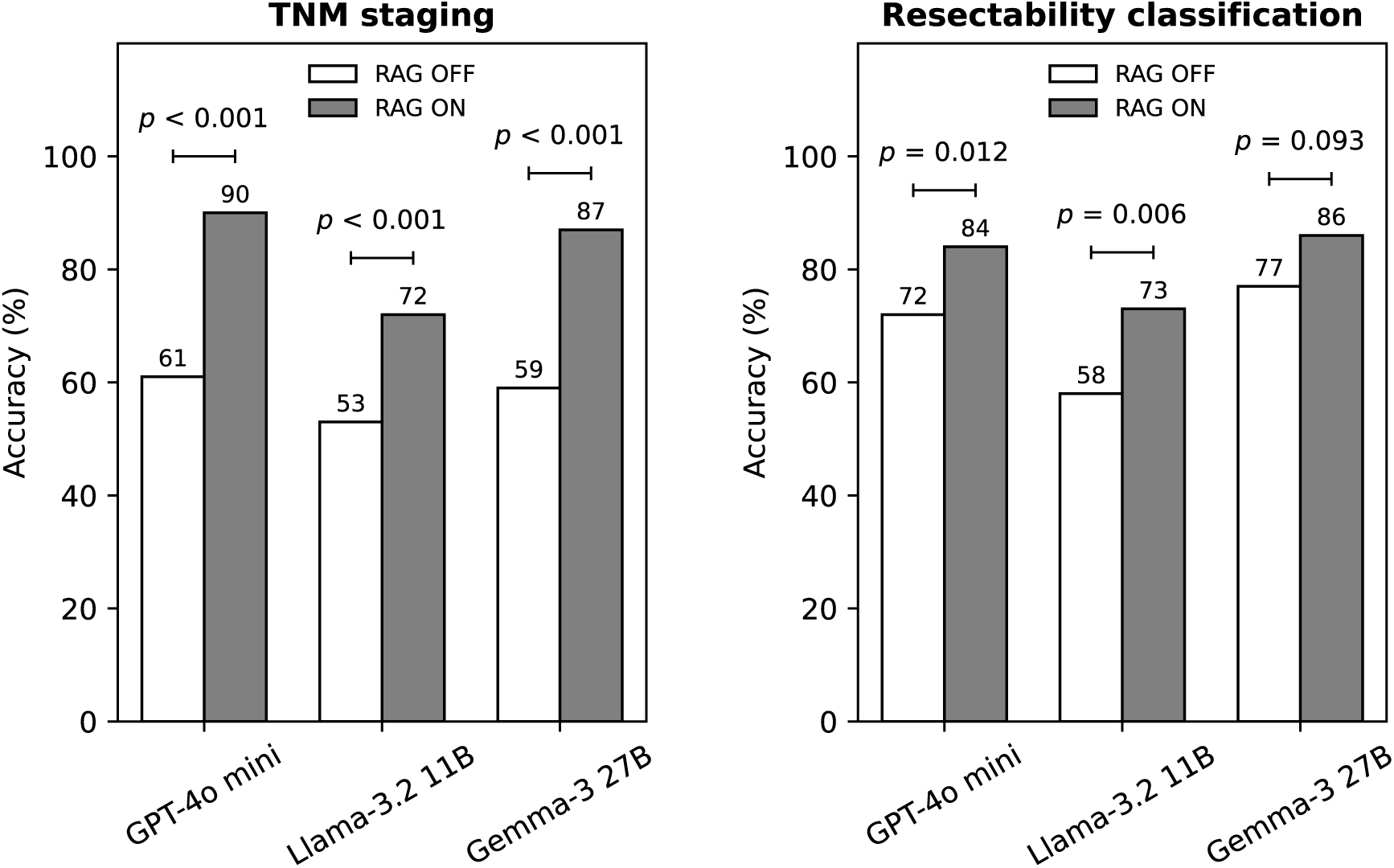
Accuracy (%) of TNM staging and resectability classification over 100 pancreatic cancer cases by our system using different LLMs (GPT-4o mini, Llama-3.2 11B, and Gemma-3 27B) with and without RAG. For each LLM, McNemar tests compared performance with and without RAG for both TNM staging and resectability classification, and the corresponding *p* values are shown. LLM = large language model, RAG = retrieval-augmented generation.

**Table 1:** Classification accuracy (%) for T, N, and M categories and local invasion among different LLMs (GPT-4o mini, Llama-3.2 11B, and Gemma-3 27B) with and without RAG.

| LLM | RAG | T category | N category | M category | Local invasion |
| --- | --- | --- | --- | --- | --- |
| GPT-4o mini | OFF | 61 | 79 | 91 | 68 |
| GPT-4o mini | ON | 86 | 84 | 97 | 80 |
| Llama-3.2 11B | OFF | 61 | 76 | 89 | 43 |
| Llama-3.2 11B | ON | 74 | 80 | 93 | 65 |
| Gemma-3 27B | OFF | 63 | 79 | 91 | 51 |
| Gemma-3 27B | ON | 82 | 81 | 97 | 79 |

Representative examples of system outputs are shown in Figure 4 and in Supplemental Figures S1 and S2. In Case 55, Gemma-3 27B without RAG produced incorrect reasoning with no guideline-based justification, leading to erroneous final answers (Figure 4; Supplemental Figure S1), whereas with RAG it generated guideline-supported and correct reasoning and produced the correct final answers (Figure 4; Supplemental Figure S2). The guideline contents of chunk [12] for N classification and chunk [16] for resectability are shown in Supplemental Figure S3. The full contents of all chunks can be accessed at https://github.com/mohehe1234/local-rag/tree/v1.0.0-with-results, and the function *f*: 2*^A^* → {0,1} introduced in the Materials and Methods section was defined by *f*^−1^(1) = *U*_+_ for local invasion, *f*^−1^(1) = *U*, for T category, *f*^−1^(1) = *U*_12_ for N category, *f*^−1^(1) = *U*_12_ ∪ *U*_13_ for M category, *f*^−1^(1) = *U*_16_ for resectability, and *f*^−1^(1) = *U*_15,_ for TNM stage, where *U_i_* = {*C* ∈ 2*^A^* | chunk [*i*] ∈ *C*}.

**Figure 4:**
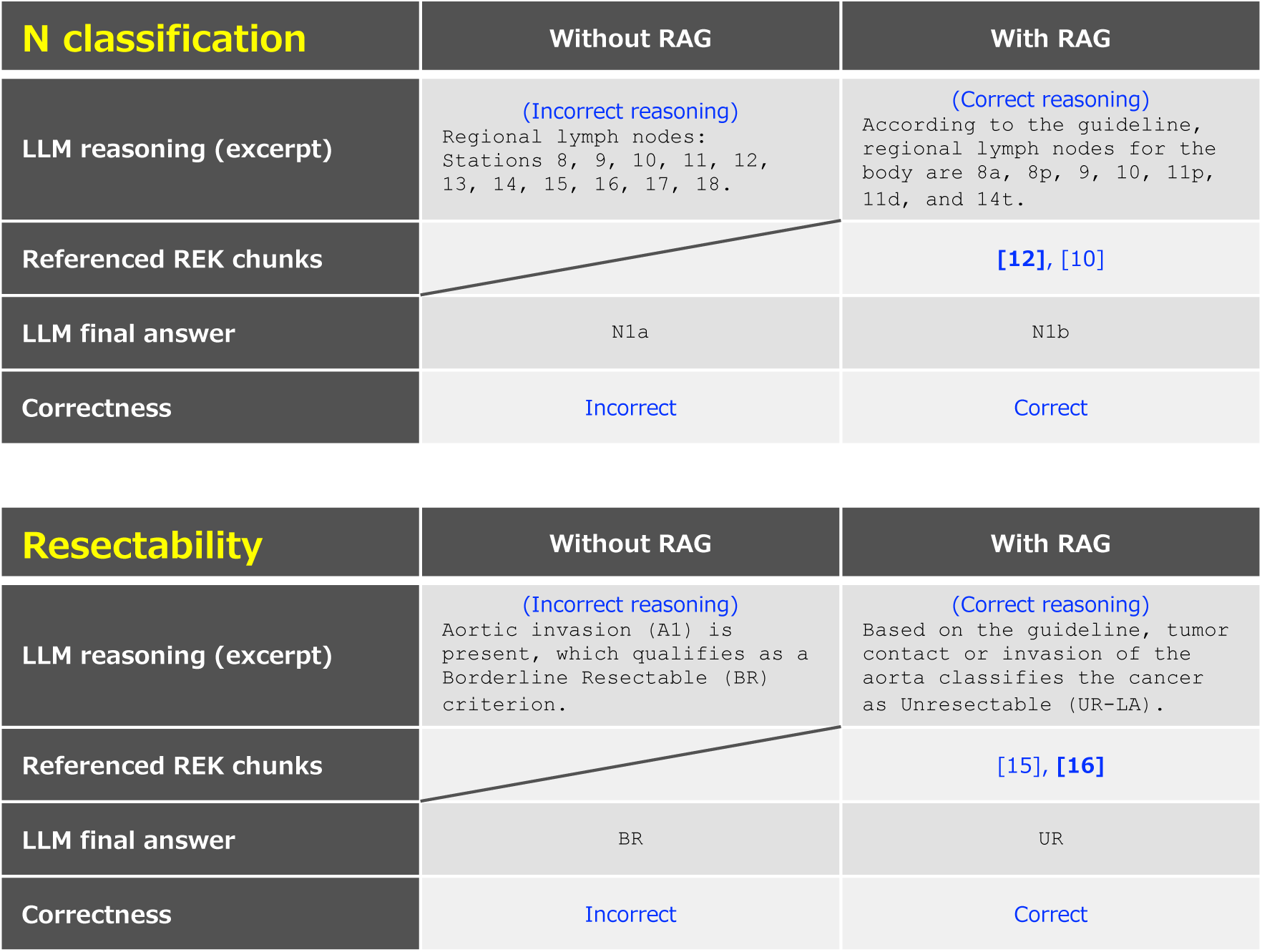
Comparison of system outputs generated without RAG and with RAG. In this example, Gemma-3 27B was used as the base LLM to perform pancreatic cancer staging for Case 55. For N classification, the model without RAG misidentified the current definition of regional lymph nodes and produced an incorrect final answer, whereas the model with RAG correctly referenced the relevant REK section (chunk [12]) and generated the correct final answer. For resectability classification, the model without RAG again exhibited incorrect reasoning and an incorrect final answer, while the model with RAG referenced the appropriate REK section (chunk [16]) and produced the correct answer. LLM = large language model, RAG = retrieval-augmented generation, REK = reliable external knowledge.

As summarized in Table 2, we calculated the mean context recall and context precision across 100 cases for each LLM and task. Context recall was 1 throughout, confirming that all required information was successfully retrieved. Context precision was 1 for most tasks with GPT-4o mini and Gemma-3 27B, indicating that the needed information was typically captured in the top chunk. For Llama-3.2 11B, context precision values of 0.5 occurred more often, but this still reflected retrieval of the required information within the top two chunks.

**Table 2:** Retrieval performance of the three RA-LLMs for each staging task, summarized by the mean context recall and mean context precision.

| LLM | Staging task | Context recall | Context precision |
| --- | --- | --- | --- |
| GPT-4o mini | Local invasion | 1 | 1 |
| GPT-4o mini | T category | 1 | 1 |
| GPT-4o mini | N category | 1 | 0.5 |
| GPT-4o mini | M category | 1 | 1 |
| GPT-4o mini | Resectability | 1 | 1 |
| GPT-4o mini | TNM stage | 1 | 1 |
| Llama-3.2 11B | Local invasion | 1 | 0.5 |
| Llama-3.2 11B | T category | 1 | 1 |
| Llama-3.2 11B | N category | 1 | 0.5 |
| Llama-3.2 11B | M category | 1 | 0.5 |
| Llama-3.2 11B | Resectability | 1 | 0.5 |
| Llama-3.2 11B | TNM stage | 1 | 1 |
| Gemma-3 27B | Local invasion | 1 | 1 |
| Gemma-3 27B | T category | 1 | 1 |
| Gemma-3 27B | N category | 1 | 1 |
| Gemma-3 27B | M category | 1 | 1 |
| Gemma-3 27B | Resectability | 1 | 0.5 |
| Gemma-3 27B | TNM stage | 1 | 1 |

The processing time for pancreatic cancer staging is summarized in Figure 5. As expected, the RAG workflow required longer execution times than the non-RAG workflow, with median execution times increasing from 4 to 41 seconds for GPT-4o mini, from 3 to 14 seconds for Llama-3.2 11B, and from 13 to 39 seconds for Gemma-3 27B. Gemma-3 27B took longer than Llama-3.2 11B, but its processing time was comparable to that of GPT-4o mini.

**Figure 5:**
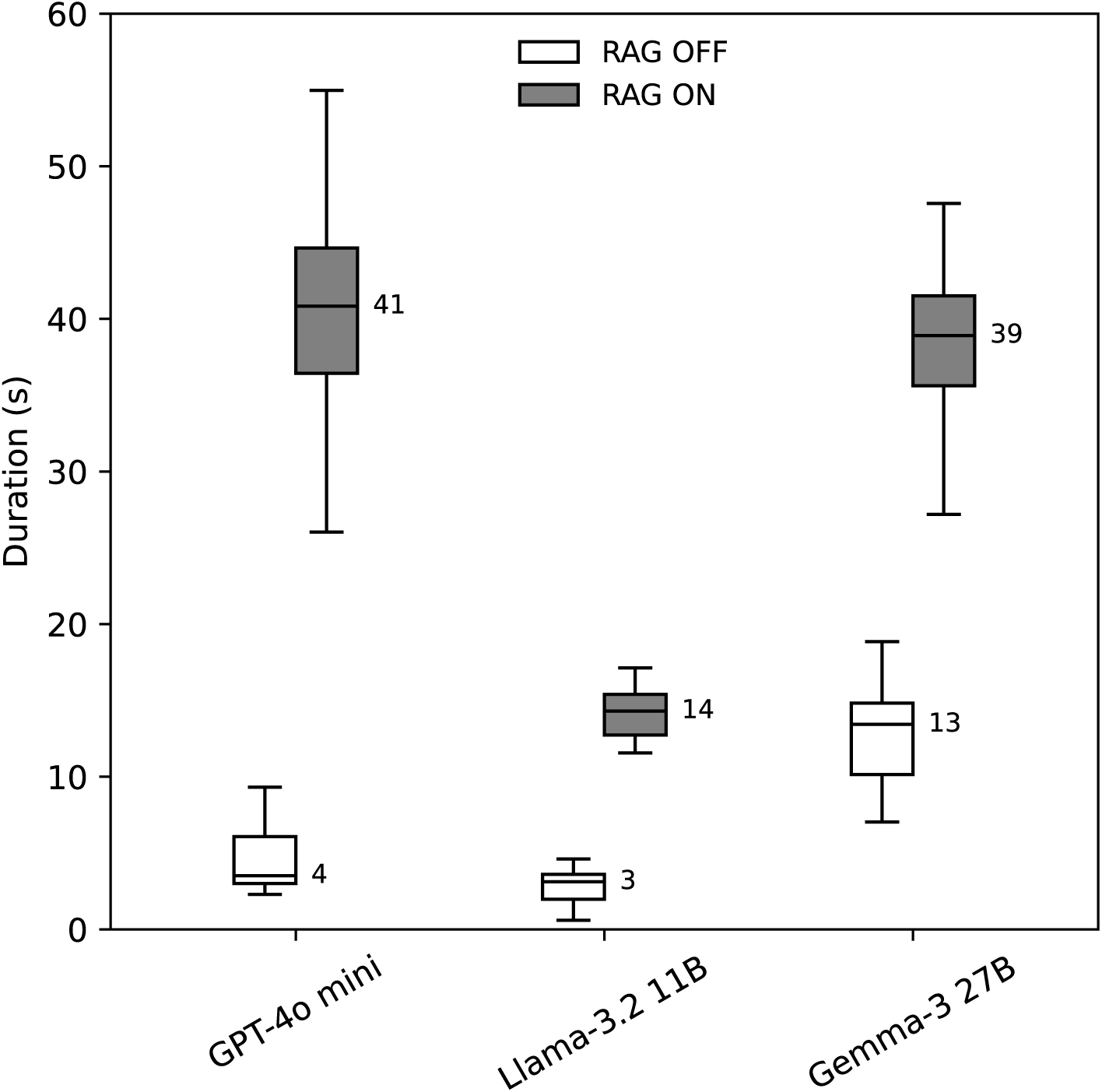
Box plot illustrating the duration (in seconds) required for pancreatic cancer staging across 100 cases among different LLMs (GPT-4o mini, Llama-3.2 11B, and Gemma-3 27B) with and without RAG. Each box represents the interquartile range (IQR), the horizontal line indicates the median, and the whiskers extend to values within 1.5 times the IQR. Outliers beyond the whisker range were omitted, and median values are shown numerically beside each box. LLM = large language model, RAG = retrieval-augmented generation.

## Discussion

Although guideline-grounded reasoning via RAG and secure offline execution are increasingly emphasized for LLMs in radiology, systems that integrate both remain scarce, as outlined in the Introduction. One reason may be that local LLMs have historically lagged far behind cloud-based models. Recent advances, however, have been remarkable; in particular, Gemma-3 27B has emerged as a high-performance local model comparable to Google’s earlier flagship cloud-based LLM, Gemini-1.5 Pro (15). Against this background, we developed an offline-deployable RAG-equipped LLM system for diagnostic radiology and publicly released its source code and validation data.

Applying our system to 100 simulated pancreatic cancer cases demonstrated that RAG functioned effectively across all tested LLMs (Llama-3.2 11B, Gemma-3 27B, and GPT-4o mini). The system achieved sufficient retrieval performance, produced guideline-grounded reasoning, and improved staging accuracy. Notably, although the local model Llama-3.2 11B showed relatively lower accuracy, the more advanced local model Gemma-3 27B achieved accuracy comparable to the widely used cloud-based GPT-4o mini, and the local models ran at speeds comparable to or faster than GPT-4o mini on our hardware. Together, these findings indicate the effectiveness and practical feasibility of our RAG system for guideline-based classification tasks.

By releasing an open-source, locally executable RAG system for diagnostic radiology, we aim to position this work as a foundation for developing clinically useful offline-deployable RA-LLMs.

Nevertheless, several limitations should be acknowledged. First, the evaluation relied on simulated cases, and real-world variability remains to be examined. Second, whether the system actually improves diagnostic accuracy or workflow efficiency when used by clinicians has not yet been assessed. Third, our system depends on a single guideline source, and its generalizability to other diseases or guideline structures has not been tested. Future work should explore broader clinical applications, such as supporting differential diagnosis, and evaluate clinical utility in real-world settings.

## Funding

This study was partially supported by JSPS KAKENHI Grant Number JP24K06686. Our department also received a scholarship grant from Guerbet Japan K.K.

## Acknowledgments

None

## Data sharing statement

Data analyzed during this study are fully available at: https://github.com/mohehe1234/local-rag/tree/v1.0.0-with-results

## Notes

### Competing Interest Statement

The authors have declared no competing interest.

### Summary of Updates

A submission-related statement was removed. No changes were made to the scientific content.

