## Supplemental Figures for "Open-Source Offline-Deployable Retrieval-Augmented Large Language Model for Assisting Pancreatic Cancer Staging"

### LLM reasoning without RAG (Case 55)

```
Okay, I will perform the requested tasks based strictly on the provided image findings and the implicit JPS
staging guideline.

**Task 1: Diagnose the local invasion factors**
Based on the findings:
*   **CH**: 0 (No mention of common hepatic artery invasion)
*   **DU**: 0 (No mention of duodenal invasion)
*   **S**: 1 (Serosal invasion noted)
*   **RP**: 1 (Retropancreatic tissue invasion noted)
*   **PV**: 0 (No mention of portal vein invasion)
*   **A**: 1 (Aortic invasion noted)
*   **PL**: 0 (No mention of Peripancreatic lymph node invasion)
*   **OO**: 0 (No mention of other organs invasion)
**Response:** CH0, DU0, S1, RP1, PV0, A1, PL0, OO0

---

**Task 2: Determine the T classification**
The tumor size is 39mm, which would normally classify as T2. However, the presence of serosal invasion (S1),
retropancreatic invasion (RP1), and aortic invasion (A1) all indicate a T3 classification. Since extension-based
criteria (T3) take precedence, the T classification is T3.
**Response:** T3

---

**Task 3: Determine the N classification**
First, identify regional lymph nodes based on tumor location (body of the pancreas):
*   Regional lymph nodes: Stations 8, 9, 10, 11, 12, 13, 14, 15, 16, 17, 18.
Next, identify metastatic regional lymph node stations:
*   Stations 9 and 10 have metastatic nodes.
Count the number of involved regional nodes:
*   Two nodes in station 9 + two nodes in station 10 = 4 involved regional nodes.
Therefore, the N classification is N1a (1–3 regional nodes involved).
**Response:** N1a

---

**Task 4: Determine the M classification**
The findings state "No other metastases are observed." This indicates no distant metastases.
**Response:** M0

---

**Task 5: Determine the resectability classification**
Based on the local invasion factors and distant metastasis:
*   Aortic invasion (A1) is present, which qualifies as a Borderline Resectable (BR) criterion.
Therefore, the resectability classification is Borderline Resectable (BR).
**Response:** BR

---

**Task 6: Determine the overall stage**
Based on the JPS staging system and the determined T, N, and M categories (T3, N1a, M0):
*   T3, N1a, M0 corresponds to **Stage III**.
**Response:** Stage III
```

### LLM final answer compared with ground truth

| | CH | DU | S | RP | PV | A | PL | OO | T | N | M | Resect. | Stage |
|---|---|---|---|---|---|---|---|---|---|---|---|---|---|
| **LLM final answer** | CH0 | DU0 | S1 | RP1 | PV0 | A1 | PL0 | OO0 | T3 | N1a | M0 | BR | III |
| **Ground truth** | CH0 | DU0 | S1 | RP1 | PV0 | A1 | PL0 | OO1 | T3 | N1b | M0 | UR | IIB |
| **Match** | ✅ | ✅ | ✅ | ✅ | ✅ | ✅ | ✅ | ❌ | ✅ | ❌ | ✅ | ❌ | ❌ |

**Supplemental Figure S1:** System output for Case 55 generated by Gemma-3 27B (as the base LLM) without RAG. The upper panel shows the full LLM reasoning output, and the lower panel presents the LLM final answer compared with the ground truth. LLM = large language model, RAG = retrieval-augmented generation.

**LLM reasoning with RAG (Case 55)**

(Referenced chunk: [6])
CH0, DU0, S1, RP1, PV0, A1, PL0, OO1

(Referenced chunks: [5], [15])
T3.
**Reasoning:**
1. **Local Invasion Factors (from Task 1):** CH0, DU0, S1, RP1, PV0, A1, PL0, OO1. This indicates serosal invasion (S1), retropancreatic tissue invasion (RP1), and contact with the aorta (A1).
2. **Tumor Size:** The tumor measures 39 mm, which is greater than 20 mm.
3. **T Category Determination:** According to the guideline, a tumor greater than 20mm *and* with extension beyond the pancreas (as indicated by serosal, retropancreatic, and aortic invasion) is classified as T3. The guideline states T3 is defined as tumor extending beyond the pancreas but not involving the CA or SMA. The patient's tumor involves the aorta (A1), but not the CA or SMA.
Therefore, the appropriate T classification is T3.

(Referenced chunks: [12], [10])
N1a
**Reasoning:**
1. **Tumor Location & Regional Lymph Nodes:** The tumor is in the body of the pancreas. According to the guideline, regional lymph nodes for the body are 8a, 8p, 9, 10, 11p, 11d, and 14t.
2. **Metastatic Nodes:** Metastases are observed in stations 9 and 10. Both stations 9 and 10 are listed as regional lymph nodes for the body of the pancreas.
3. **Counting Metastatic Nodes:** There are two metastatic nodes in station 9 and two in station 10, for a total of four metastatic regional lymph nodes.
4. **N Classification:** According to the guideline, N1a = 1-3 regional lymph nodes, N1b = 4 or more. Since there are four metastatic regional lymph nodes, the N classification is N1b.
**Correction:** My apologies. I misread the N1a/N1b criteria. The correct N classification is **N1b** due to the presence of four metastatic regional lymph nodes.

(Referenced chunks: [12], [5])
M0
**Reasoning:**
The provided guideline explicitly states that metastasis found in *non-regional* lymph nodes constitutes M1.
The patient information details lymph node metastases in stations 9 and 10. According to the guideline and the patient's tumor location (body), stations 9 and 10 *are* regional lymph nodes.
Therefore, the presence of metastases within regional lymph nodes does *not* qualify as distant metastasis (M1).
Since no distant metastases are reported, the M classification is M0.

(Referenced chunks: [15], [16])
UR-LA
**Reasoning:**
1. **T Classification (from Task 2):** T3
2. **N Classification (from Task 3):** N1b
3. **M Classification (from Task 4):** M0
4. **Local Invasion:** The patient has tumor contact/invasion of the aorta (identified in Task 1 and patient information).
Based on the guideline, tumor contact or invasion of the aorta classifies the cancer as Unresectable (UR-LA).
The presence of regional lymph node involvement (N1b) does not change this classification, as it is not considered distant metastasis.
Therefore, the resectability classification is UR-LA.

(Referenced chunks: [15], [2])
Based on the provided information and guideline:
• **T classification:** T3 (from Task 2)
• **N classification:** N1b (from Task 3)
• **M classification:** M0 (from Task 4)
Referring to Table 3 in the guideline, a T3 tumor with N1b and M0 corresponds to **Stage IIB**.

**LLM final answer compared with ground truth**

| | CH | DU | S | RP | PV | A | PL | OO | T | N | M | Resect. | Stage |
|---|---|---|---|---|---|---|---|---|---|---|---|---|---|
| LLM final answer | CH0 | DU0 | S1 | RP1 | PV0 | A1 | PL0 | OO1 | T3 | N1b | M0 | UR | IIB |
| Ground truth | CH0 | DU0 | S1 | RP1 | PV0 | A1 | PL0 | OO1 | T3 | N1b | M0 | UR | IIB |
| Match | ✅ | ✅ | ✅ | ✅ | ✅ | ✅ | ✅ | ✅ | ✅ | ✅ | ✅ | ✅ | ✅ |

**Supplemental Figure S2:** System output for Case 55 generated by Gemma-3 27B (as the base LLM) with RAG. The upper panel shows the full LLM reasoning output, and the lower panel presents the LLM final answer compared with the ground truth. LLM = large language model, RAG = retrieval-augmented generation.

**Example retrieved chunks from REK**

```
(Chunk [12])
### Regional lymph nodes
Regional lymph nodes are not synonymous with lymph nodes to be dissected. If metastasis is found in a lymph node
other than a regional lymph node, it is defined as M1 (Table 2). In the pancreatic head and body, lymph node 14
(tumor side; 14t) is considered a regional lymph node; however, if metastasis is found in lymph node 14 (tumor
opposite side; 14op), it is not considered M1 but instead warrants further investigation in future studies.

Table 2. Regional lymph nodes.
| | Head | Body | Tail |
|--|--|--|--|
| Regional lymph nodes | 6, 8a, 8p, 12a, 12b, 12p, 13, 14t, 17 | 8a, 8p, 9, 10, 11p, 11d, 14t, 18 | 8a, 9, 10,
11p, 11d, 18 |

Note: Reproduced with permission from the Japan Pancreas Society and Kanehara & Co., Ltd.: Japanese
Classification of Pancreatic Carcinoma, eighth edition, 2023.

### Recording lymph node metastases
In resected cases, the number of dissected and metastatic nodes should be recorded for each lymph node.

- NX: Regional lymph nodes cannot be assessed
- N0: No regional lymph node metastasis
- N1: Regional lymph node metastasis
  - N1a: Metastasis in one to three regional lymph nodes
  - N1b: Metastasis in four or more regional lymph nodes

## Distant metastases (M category)
- M0: No distant metastasis
- M1: Distant metastasis

The M1 site should be recorded using the following codes:


(Chunk [16])
## Resectability classification
### Resectable: R
No tumor contact with the superior mesenteric vein (SMV) or portal vein (PV), less than 180 contacts, or invasion
without occlusion. The clear fat planes around the SMA, CA, and common hepatic artery (CHA) show no contact or
invasion.

### Borderline resectable: BR
Subclassified according to SMV/PV invasion alone or arterial invasion.
- BR-PV (SMV/PV invasion alone): No findings of contact or invasion of the SMA, CA, or CHA. Tumor contact or
invasion of the SMV/PV of 180°degrees or more, or occlusion of the SMV/PV, not exceeding the inferior border of
the duodenum.
- BR-A (arterial invasion): Tumor contact or invasion of the SMA and/or CA of less than 180 degrees without
stenosis or deformity. Tumor contact or invasion of the CHA without tumor contact or invasion of the PHA and/or
CA.

### Unresectable: UR
Subclassified according to the status of distant metastasis.
- UR-LA (locally advanced): tumor contact or invasion of the SMV/PV of 180° or more, or occlusion of the SMV/PV
extending beyond the inferior border of the duodenum. Tumor contact or invasion of the SMA and/or CA of 180°or
more. Tumor contact or invasion of the CHA, showing tumor contact or invasion of the PHA and/or CA. Tumor contact
or invasion of the aorta.
- UR-M (tumor with distant metastasis): distant metastasis, including nonregional lymph node metastasis.

# Surgical treatment
## Types of operative procedures
### Surgery with or without resection
```

**Supplemental Figure S3:** Example retrieved chunks from the REK. Chunk [12] contains information required for N classification, and chunk [16] provides information necessary for resectability classification. REK = reliable external knowledge.

This material partially reproduces content from Ishida M et al. (2024), Japanese classification of pancreatic carcinoma, Eighth edition, J Hepato-Biliary-Pancreat Sci, 31:755–768. © 2024 The Author(s), published under the CC BY 4.0 license.
